# Safety and immunogenicity of COReNAPCIN^®^, a SARS-CoV-2 mRNA vaccine; a randomized, double-blind, placebo-controlled phase 1 clinical trial

**DOI:** 10.1101/2023.11.01.23297898

**Authors:** Mohammadreza Salehi, Ilad Alavi Darazam, Alireza Nematollahi, Masoumeh Alimohammadi, Sedigheh Pouya, Reza Alimohammadi, Nasim Khajavirad, Meysam Porgoo, Mosslim Sedghi, Mohammad Mahdi Sepahi, Maryam Azimi, Hamed Hosseini, Seyed Mahmoud Hashemi, Somaye Dehghanizadeh, Vahid Khoddami

## Abstract

The repeated COVID-19 outbreaks, despite global vaccination, highlights the need for booster doses. Here, we present the outcomes, up until day 90 post vaccination, of a randomized, double-blind, placebo-controlled phase 1 clinical trial of the mRNA-based vaccine candidate; COReNAPCIN^®^, as a booster dose in adults aged 18-50 who had previously received three doses of inactivated vaccines. In the study, 30 participants randomly (2:2:1) received 25 μg, or 50 μg of COReNAPCIN^®^, or placebo. The results indicated that COReNAPCIN^®^ was well tolerated in vaccinated individuals in both groups with no life-threatening or other serious adverse events. The most noticeable solicited adverse events were pain at the site of injection, fatigue and myalgia. Regarding the immunogenicity, given the seroprevalence of SARS-CoV-2 antibodies due to the vaccination history for all, and previous SARS-CoV-2 infection for some participants, the recipients of COReNAPCIN^®^, two weeks post vaccination, showed significant fold increases in the level of anti-RBD (6.6 and 8.1 folds) or anti-spike (11.5 and 21.7 folds), and potent neutralizing antibodies (10.2 and 8.4 folds) in 25 and 50 μg groups, respectively, while no meaningful changes were observed in the placebo group. Additionally, the significant increase in the spike-specific IFN-γ T-cell response upon vaccination, underscores the activation of cellular immunity. Altogether, the favorable safety, tolerability, and immunogenicity profile of COReNAPCIN^®^ support its further clinical development.

**Trial registration number:** **IRCT20230131057293N1**

## Introduction

The ongoing COVID-19 pandemic, caused by the severe acute respiratory syndrome coronavirus 2 (SARS-CoV-2), persists due to the evolution of immune-escaping new strains, and waning immunity in vaccinated individuals - with the global symptomatic or asymptomatic spread of infection^1,2.^ Therefore, developing safe and effective COVID-19 vaccines as booster doses, preferentially against specific strains, is still a requirement for protecting lives and reducing healthcare costs^3^. Heterologous vaccination schedules, characterized by the administration of diverse vaccine types for primary and booster doses, have exhibited promising outcomes in eliciting robust and long-lasting immune responses^4-9^. Among various vaccination approaches, mRNA-based vaccines have emerged as a leading solution to prevent COVID-19 infection more effectively^10-14^.

We have previously described the pre-clinical development of COReNAPCIN^®^, a nucleoside modified mRNA-based vaccine formulated in lipid nanoparticles (LNPs) against the original (Wuhan) SARS-CoV-2 Strain^15^.

In this report, we present the phase 1 clinical study that aimed to assess the safety, tolerability, and immunogenicity of COReNAPCIN^®^ in humans when used as a booster dose in adults aged 18-50 who had previously received three doses of inactivated vaccines. This study setting was advised by Iranian Food and Drug Administration (IFDA) as majority of vaccinated individuals in Iran at the time of conducting this clinical trial had the vaccination history of three doses of inactivated vaccines (either Sinopharm^®^ or BIV1-CovIran^®^). The study was conducted from February to May 2023 and the outcomes are presented here up until day 90 post vaccination.

The objective of this study was primarily safety evaluation, followed by immunogenicity assessment to gain insights into the potential of COReNAPCIN^®^ administration to reinforce protection against SARS-CoV-2 infection. Importantly, as updated COReNAPCIN^®^ against new variants (including Omicron XBB.1.5, XBB.1.16 & EG.5.1) is currently under development, the findings from phase 1 clinical trial of the vaccine against the original Wuhan strain carry significant implications for public health strategists, especially regarding the safety concerns, to focus on the immunogenicity potential of new strain-specific COReNAPCIN^®^ vaccine candidate as booster dose.

## Results

### Trial Participants

Between February and May 2023, a total of 93 healthy individuals were screened, out of them 30 participants were randomized into three groups; twelve participants in group 1 received 25 μg COReNAPCIN^®^, twelve participants in group 2 received 50 μg COReNAPCIN^®^, and six participants in placebo group received normal saline (**Fig. 1**). Table 1 illustrates an overview of the demographic attributes based on the treatment allocation.

**Table 1.**
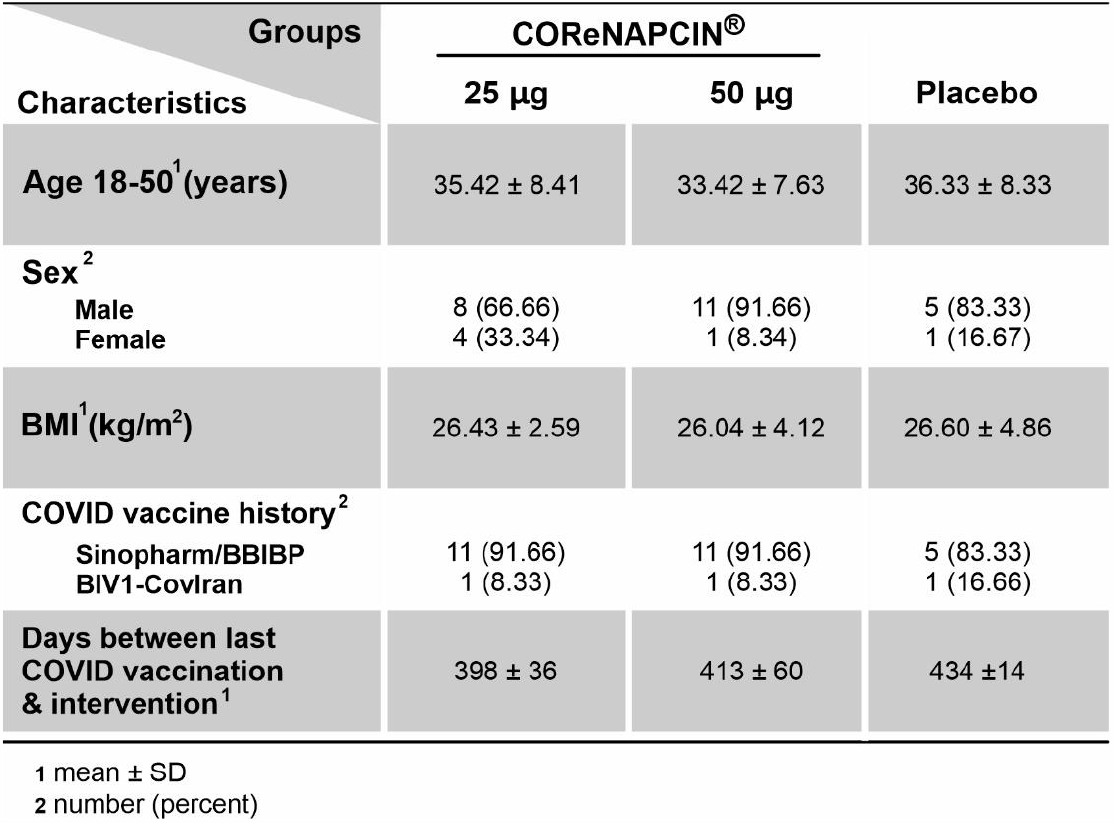
Overview of demographics for study intervention recipients.

**Figure 1.**
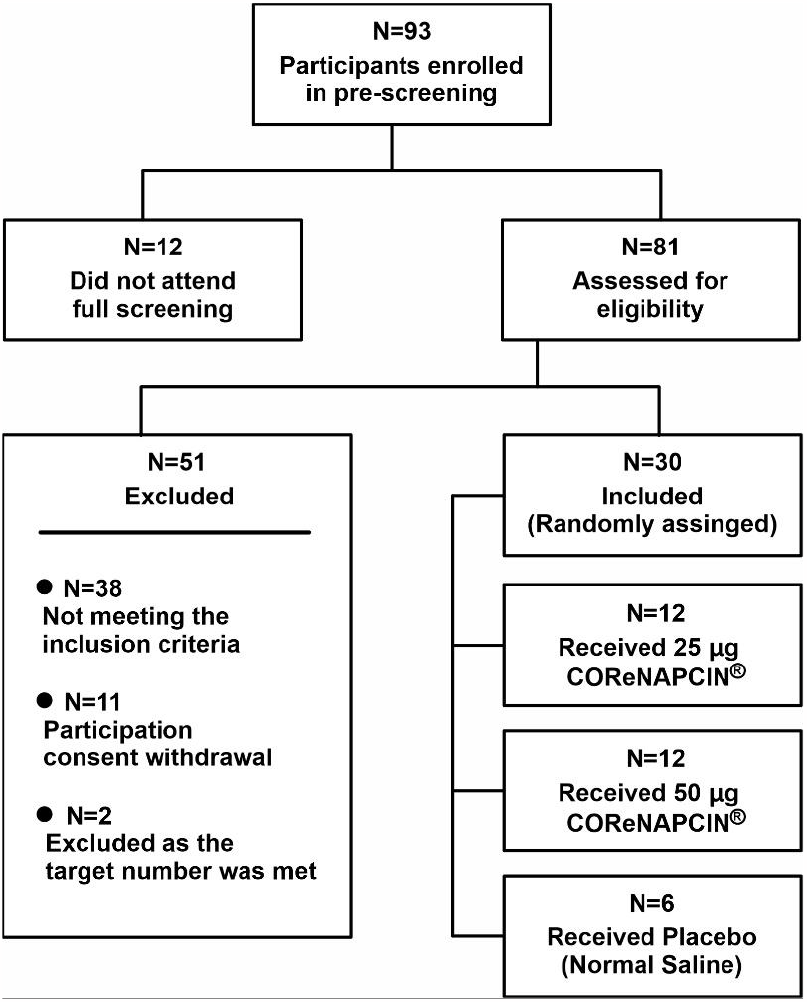
Trial participant recruitment and randomization. The safety population included all participants who received injection of the COReNAPCIN^®^ vaccine or the placebo.

## Safety

A comprehensive overview of adverse events (AEs) experienced by participants during the study is provided in figure 2, mapping local and systemic AEs. As well as good tolerability, COReNAPCIN^®^ demonstrated lack of serious effects at both implemented doses of 25 μg and 50 μg, resulting in no reported serious AEs, study discontinuation, events of special interest, new chronic diseases, or potential immune-mediated medical conditions.

**Figure 2.**
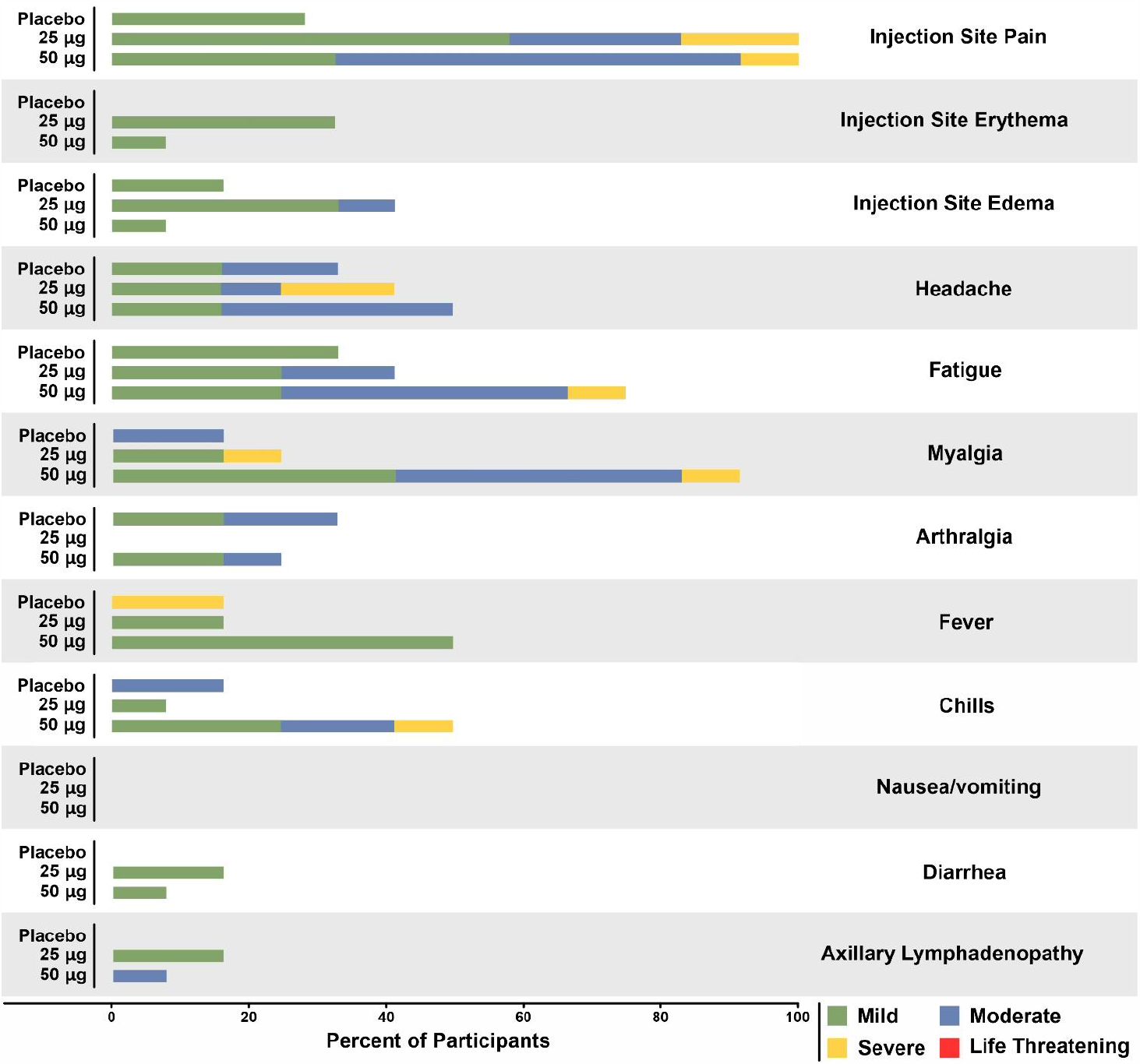
The solicited local and systemic adverse events. The graph shows the percentage of participants in each vaccination group (25 μg, 50 μg, and placebo) who experienced solicited local and systemic adverse events. These events are graded according to the FDA’s Toxicity Grading Scale for vaccines, which categorizes them as Grade 1 (mild), Grade 2 (moderate), Grade 3 (severe), or Grade 4 (life-threatening).

Most participants reported only mild or moderate, local or systemic solicited reactions following the injection (**Fig. 2**, more detail in Supplementary Table 1). In 25 μg group, severe solicited systemic reactions, including headache and myalgia, were reported by two participants (16.7%) (**Fig. 2**), while two participants (16.7%) experienced severe injection site pain as a solicited local reactions (**Fig. 2**). In 50 μg group, three participants (25.0%) reported severe solicited systemic reactions, such as myalgia, chills, and fatigue (**Fig. 2**), and one participant (8.3%) reported severe solicited local reactions in the form of injection site pain (**Fig. 2**).

Overall, the 50 μg dose group showed a higher incidence rate of solicited local and systemic reactions. The most frequently reported solicited local event was pain at the injection site. Other systemic adverse events were headache, fatigue, and myalgia.

Unsolicited AEs are summarized in supplementary Table 2. Among the severe unsolicited AEs reported by COReNAPCIN^®^ vaccine recipients, common cold was observed in one participant in 25 μg group, which resolved in severity within a span of three days and was considered unlikely to be related to vaccination. Within 50 μg group, a single participant experienced gastrointestinal disorders 15 days after the administration of COReNAPCIN^®^ which was unlikely to be linked to the intervention, and it resolved after three days. Additionally, two severe events were also reported by participants who received the placebo. Interestingly, one case of COVID-19 infection was reported during the entire observation period in the placebo group diagnosed at day 7 after intervention and lasted until week 2. Throughout the course of the study, there were no significant clinical deviation from the normal ranges for any of the hematological or biochemical parameters tested, implying the absence of any significant medical concern.

For evaluating the possible vaccination-related immunopathologic events, release of inflammatory cytokines into the serum was assessed and the results indicated no substantial increase associated with hyperactivation of the immune system.

## Immunogenicity

### Anti-Spike, Anti-RBD and Neutralizing Antibodies Response

COReNAPCIN^®^ administration led to significant rise in anti-Spike (anti-S) and anti-RBD IgG antibody levels within 15 days in the administered groups. The subset of individuals who were given a 50 μg dose exhibited a remarkable 21.7-fold increase in binding anti-spike antibodies (with a highly significant P-value of < 0.0001). Similarly, in the 25 μg dose group, there was a substantial enhancement of anti-Spike titers by a factor of 11.5 (with a P-value of < 0.001) 15 days after administration of COReNAPCIN^®^. Furthermore, the RBD antibody’s geometric mean titer (GMT) also displayed a noteworthy increase. Specifically, for individuals receiving a 50 μg dose, the GMT of the RBD antibody surged by a factor of 8.1, while a slightly lower increase of 6.6 was observed in those administered a 25 μg dose. These findings are visually represented in Figures 4a and 4b, illustrating the robustness of immune responses across both dosage strengths. The figures also depict a gradual decline in antibody concentration over time, consistent with typical immune response patterns.

**Figure 3.**
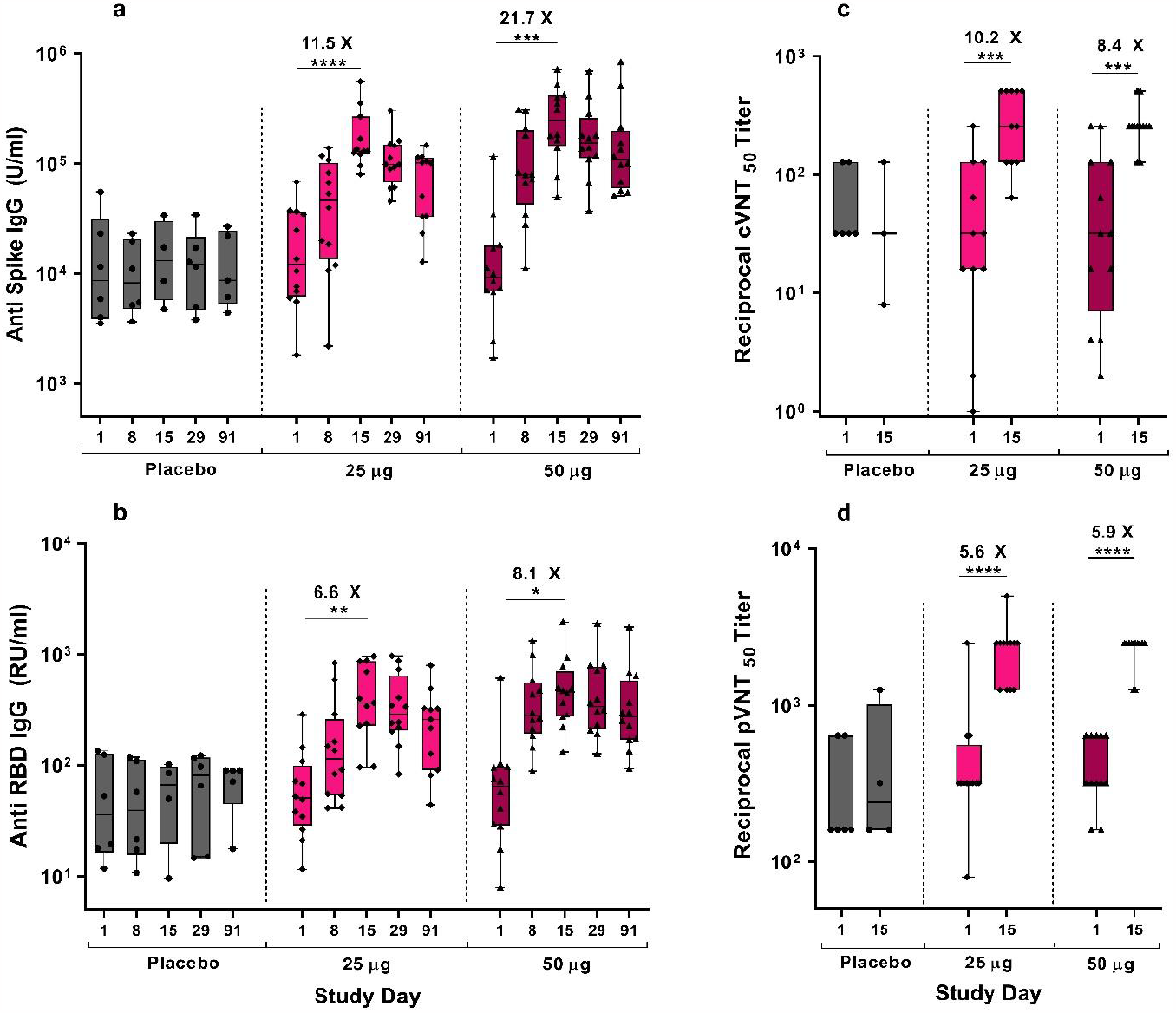
COReNAPCIN^®^ induced antibody responses. Serum samples were collected on pre- and at different days post vaccination. Using ELISA, anti-spike (a) and anti-RBD (b) binding antibody titers were measured on study days 1, 8, 15, 29, 91. The neutralizing capacity against Wuhan in the sera of study day 15 were assessed comparing to the baseline level by cVNT (c) and pVNT(d). Each data point corresponds to an individual participant. The intervention groups are categorized as follows: 25 μg, 50 μg, and placebo. The box plots display Min to Max of data. Statistical significance was defined as a P-value less than 0.05 (*P <0.05, **P <0.01, ***P <0.001, ****P <0.0001).

**Figure 4.**
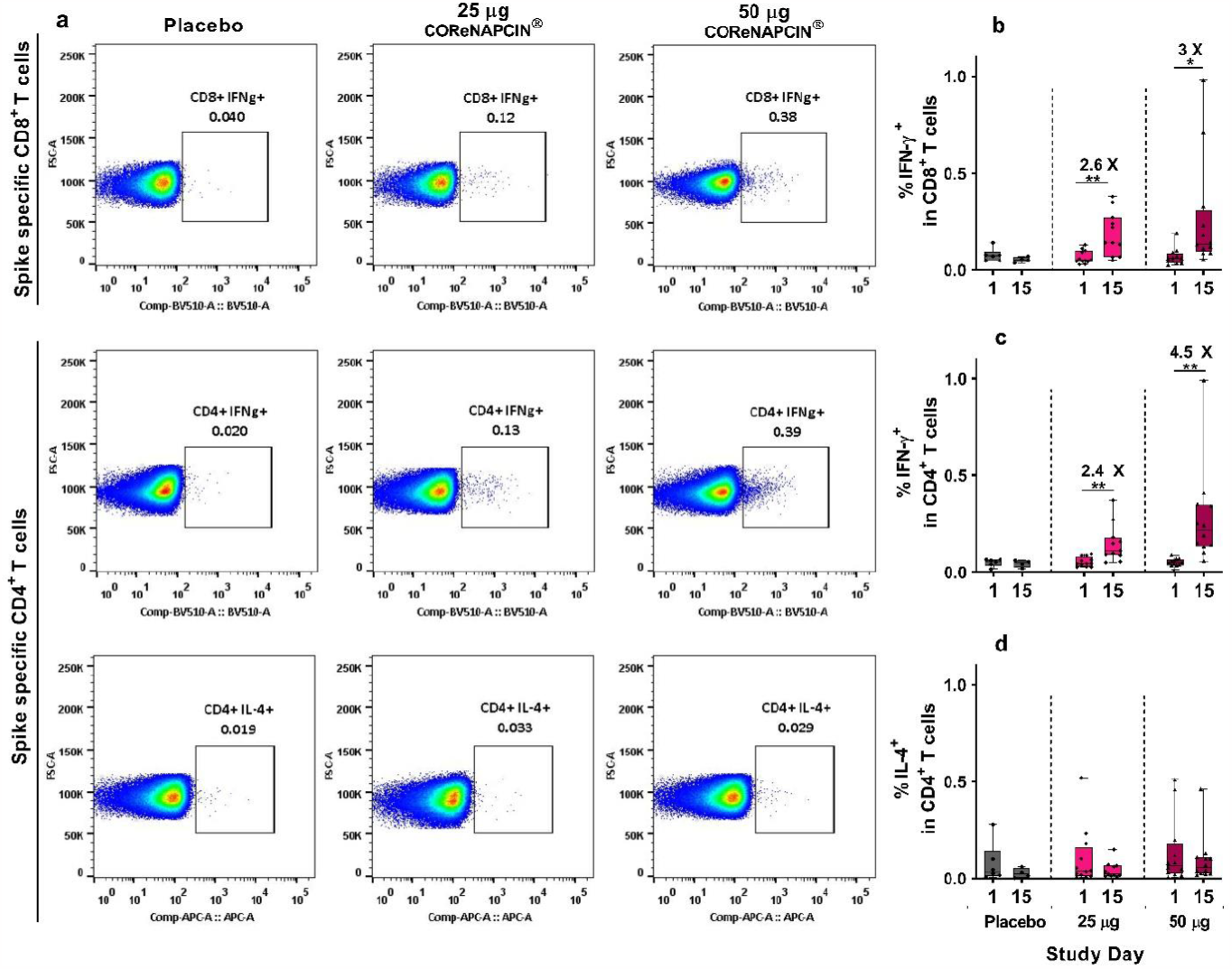
COReNAPCIN^®^ induced T-Cell responses. PBMCs from all vaccinated individuals were collected on pre-and day 14 post vaccination (study day 15). After being stimulated with SARS-CoV-2 Spike peptide pools for eighteen hours, flow cytometry was used to identify, the IFN-γ and IL-4 producing T cells (Supplementary Fig. 1 shows the gating strategy). a, Representative pseudocolor flow cytometry plots of cytokine-producing CD4^+^ and CD8^+^ T cells from 3 different study groups are shown. The percentage of spike specific IFN γ^+^ in CD8^+^ T cells (b), and IFN γ^+^ (c) and IL-4 ^+^ (d) in CD4^+^ T cell population are displayed on study day 1 and on study day 15. Each data point corresponds to an individual participant. The intervention groups are categorized as follows: 25 μg, 50 μg, and placebo. The box plots display Min to Max of data. A P-value less than 0.05 (*P <0.05, **P <0.01) indicates statistical significance.

Prior to and after vaccination, pseudovirus-based neutralization (pVN) as well as conventional virus neutralization (cVN) tests were conducted in order to assess neutralization activity. The COReNAPCIN^®^ vaccinated groups exhibited neutralizing antibody IC_50_ reciprocal titers (measured using cVN test) that were as a minimum of eight times higher than those of the control group (**Fig. 4c**). In case of pVN antibody levels, in the sera of all participants in both groups, at least 2-fold increase in neutralization activity on day 15 study was observed (**Fig. 4d**). As a result of the vaccination, pVN antibodies levels for both strengths increased by approximately 5.5-fold after the vaccination; nevertheless, there was no statistically significant difference between the subjects in 25 μg and 50 μg groups (P < 0.0001) **(Fig. 4d**).

### SARS-CoV-2 T-cell Response

B cell epitopes, which are components of antigens that the immune system recognizes, can mutate rapidly. Because of this, cellular immunity plays a crucial role in protecting against these mutating epitopes. Herein, we studied antigen-specific T cells that were induced by a booster dose of COReNAPCIN^®^. We collected blood samples on study day 1 and 15 and stimulated the T cells with spike peptide pool. Using flowcytometry method, we measured the percentage of T cells producing the cytokines IFN-γ and IL-4 (**Fig. 5**).

In most of the COReNAPCIN^®^ vaccinated participants, the frequency of IFN-γ^+^ CD8^+^ T cells were significantly increased comparing to their baseline values, with a P-value of < 0.01 and < 0.05 for 25 μg and 50 μg groups, respectively (**Fig. 5b**). We also assessed the IFN-γ induction in CD4^+^ T cells and the results showed a 4.5-fold increase (p< 0.01) in the 50 μg dose group at day 14 after vaccination (study day 15) comparing the pre vaccination level. This value was increased by a factor of 2.4 in 25 μg COReNAPCIN^®^ vaccinated group (p< 0.01) (**Fig. 5c**). Additionally, the IFN-γ response in CD8^+^ and CD4^+^ population was substantially higher than that of placebo group demonstrating a strong induced cellular immune response. In order to determine the polarization of T cell responses, we assessed the IL-4 secretion in CD4^+^ T cell population following re-stimulation with spike peptide pools. In both of the dose groups, the vaccine did not elicit IL-4^+^ CD4^+^ T cells (**Fig. 5d**). These results accord with preclinical evidence, suggest that COReNAPCIN^®^ does not bias towards a Th2 immune response which rule out the potential occurrence of Vaccine-Associated Enhanced Respiratory Disease (VARED).

## Discussion

Despite extensive efforts and strict regulations and restrictions in different countries over the past three years, and global consensus to eradicate, or prevent the spread of SARS-CoV-2 virus, the world still struggles with the disease. Undoubtedly, the most effective solution is vaccination; most likely annually and preferentially with strain-specific updated mRNA booster doses^16-18^. In Iran, several COVID-19 vaccines, usually with inactived and/or recombinant platforms, have been developed ^19-24^ but according to our knowledge, COReNAPCIN^®^ is the first Iranian mRNA vaccine that has been able to enter the human clinical trial.

COReNAPCIN^®^, initially designed against the original (Wuhan) strain of SARS-CoV-2^15^, showed a favorable safety and tolerability profile in clinical setting up to a tested dose of 50 μg, with robust activation of both humoral and cellular immunity in vaccinated individuals. Individuals vaccinated with either of 25 or 50 μg doses did not experience any life-threatening condition or other serious AEs. Among the 30 participants who received COReNAPCIN^®^, the incidence rate of severe solicited adverse events was 20%, where the most common AE observed in both 25 or 50 μg doses, was pain at the site of injection, often upon touch, a common symptom observed in similar mRNA-based vaccines^25-30^. This symptom is expected and is apparently due to activation of the immune system and recruitment of the immune cells to the site of injection where muscle cells, in their cytoplasm, translate the code of COReNAPCIN^®^ mRNA into spike glycoprotein, eventually sending a trimeric, fully folded and glycosylated antigen to the cell surface to be introduced to, and activate, the immune system^31^.

Systemic side effects of mRNA-based COVID-19 vaccines mainly include mild to moderate headache, fatigue, myalgia, and chills, which are dose-related and usually disappear within 24 to 48 hours of onset^2,32^. Similarly, appearance of primarily a mild to moderate fatigue and myalgia especially upon administration of a higher COReNAPCIN^®^ dose (50 μg) is likely due to the activated immune system in a dose-dependent manner, which resolved after few days as expected without imposing any concerning health complication. The other observed transient unsolicited AEs which randomly occurred in different groups, including the placebo group, rule out any direct connection to vaccination.

COReNAPCIN^®^ administration also triggered a strong humoral immunity in both 25 or 50 μg doses as it is evident in sharp rise of anti-RBD and anti-spike binding antibodies upon vaccination. Here, it is important to consider the seroprevalence of SARS-CoV-2 antibodies in participants in this study due to vaccination history for all participants who had the record of receiving three doses of an inactivated vaccine, and some of them also had the history of previous SARS-CoV-2 infection. Therefore, the anti-RBD and anti-spike antibodies were already noticeable at the day one for all participants. Here, administration of a single dose of COReNAPCIN^®^ as booster, again in both 25 or 50 μg doses, significantly increased both of binding and neutralizing antibody levels in comparison to the unchanged states in participants of the placebo group. It is notable to mention that at the time-period of running this experiment in Iran, the country was in the rise of the 8^th^ peak of COVID-19 outbreak, since the beginning of the pandemic, affecting a large number of people most probably with circulating XBB subvariants. Among the participants in this study, during the outbreak, one COVID-19 infection was reported and in a person who was among the placebo group, but no infection was reported in individuals vaccinated with COReNAPCIN^®^.

COReNAPCIN^®^ also induced a strong cellular immune response, confirmed by IFN-γ secretion by CD4^+^ and CD8^+^ T-cells after restimulation with SARS-CoV-2 specific peptide. Activation of this arm of the immune system plays a crucial role especially against new and upcoming variants, as the majority of the spike-glycoprotein sequence is shared between new variants and their ancestors.

Overall, conducting a randomized, double-blind, placebo-controlled phase 1 clinical trial of COReNAPCIN^®^ as booster in adults previously immunized with inactivated vaccines, proved a favorable safety, tolerability, and immunogenicity profile. Especially as new variants are rapidly evolving and due to the power of mRNA technology in rapid design and production of updated vaccines, COReNAPCIN^®^ has been designed and produced for several variants of concern (VOCs) and are currently under evaluation (unpublished). Here, the outcomes of the present study support further clinical development of strain-specific COReNAPCIN^®^ for future use.

## Methods

### Trial Design, Participants, and Study Approval

In this investigation, a total of 30 participants, 18–50 years old, who had all received three doses of inactivated vaccines (Sinopharm^®^/BBIBP COVID-19 vaccine or BIV1-CovIran vaccine), were enrolled. It was imperative to meticulously document their vaccination history, which encompassed details such as the specific vaccine type, the number of doses administered, the intervals between doses, and the number of days elapsed since the most recent dose (between 3 to 18 months). Moreover, participants were required to provide written informed consent prior to their inclusion in the study. The enrolment, injections, and monitoring took place at Imam Khomeini Hospital Complex (Mahdi Clinic), Tehran, Iran, and the trial was randomized, parallel-designed, placebo-controlled, and double-blind. Its primary objective focused on evaluating both the safety and immunogenicity of different dosages of COReNAPCIN^®^.

The study comprised two cohorts, each comprising 15 participants, who were assigned to receive either doses of COReNAPCIN^®^ at 25 μg or 50 μg. The placebo group received a saline solution of 0.9%. Within each dose group, a sentinel subgroup of five participants was randomly allocated in a 4:1 ratio (vaccine: placebo) to provide initial safety data. After evaluating the safety data from the sentinel group by the data and safety monitoring board, the trial progressed to a non-sentinel expansion phase. This expansion phase included 10 participants in each group, randomized in an 8:2 ratio (vaccine: placebo). Dose 25 μg were administered in the first cohort; the study progressed to the 50 μg dose after evaluation of safety data by the data and safety monitoring board. An unblinded statistician prepared a paper list of randomization codes produced by a computer program. Participants and other study personnel (principal investigator, site coordinator, site staff) were blinded, except for staff involved in preparation of doses.

The clinical trial was a collaborative effort between the sponsor, ReNAP Therapeutics, and the contract research organizations, Pharmed Pajoohan Viera, who jointly designed the study. Their responsibilities encompassed data collection, analysis, interpretation, and report writing. Clinical monitoring, pharmacovigilance, and data management were entrusted to the Contract Research Organization, Pharmed Pajoohan Viera. To ensure the safety and progress of the trial, a safety review committee was formed from Clinical Trial Center of Tehran University of Medical Sciences and an independent data and safety monitoring board (DSMB). This committee diligently assessed safety data and closely monitored the overall advancement of the trial, including decisions pertaining to dose escalation. Ethical compliance was upheld through the approval obtained from the Iran National Committee for Ethics in Biomedical Research (IR.NREC.1401.007). Furthermore, the trial’s registration details were duly recorded on irct.ir (IRCT20230131057293N1). The study was carried out in accordance with the Declaration of Helsinki, and Good Clinical Practice.

### Trial Vaccine and Placebo

The COReNAPCIN^®^ mRNA vaccine, developed by ReNAP Therapeutics, was provided in 2 mL Type I glass vials as a sterile, frozen, aqueous formulation with a concentration of 0.1 mg/mL. The vials were stored in a frozen state at a temperature of -20°C (with a 5°C tolerance). When thawed, the vaccine appeared as a white to off-white liquid with a nominal pH value of 7.4 and an osmolality of approximately 325 mOsm/kg. The vaccine contained mRNA encoding the complete S protein of SARS-CoV-2, formulated within lipid nanoparticles as the final pharmaceutical product. As for the placebo, it consisted of an injectable saline 0.9% solution. The vaccine or placebo were injected into the deltoid muscle of the upper arm using a 25G needle.

### Safety Assessments

Throughout the duration of the study, both solicited and unsolicited side events were recorded (Supplementary Tables 1,2). The principal investigator conducted a comprehensive evaluation of the severity, intensity, and correlation of each event with the administration of the vaccine. Following each immunization within the sentinel group, vital signs and injection site reactions were monitored for 48 hours, while the non-sentinel group underwent a monitoring period of two hours. Participants received training to record local and systemic adverse events (AEs) on a daily basis in a symptom diary for a period of 7 days. During the study visits, the site staff conducted reviews of these symptom diaries. Additionally, unsolicited events were documented and recorded during all visits and for the entirety of each participant’s engagement in the study. To ensure safety oversight, an independent group of experts (Data and Safety Monitoring Board), which advises the Iran FDA, performed thorough safety reviews.

### Enzyme-Linked Immunosorbent Assay (ELISA)

#### SARS-CoV-2 anti RBD IgG

The SARS-COV-2 RBD precoated plates (Pishtazteb, Lot: 14003) were utilized in accordance with the manufacturer’s instructions. Briefly, 100 μL of diluted serum samples, standards or control solutions pipetted into the wells and incubated at 37°C for half an hour. Subsequently, the plates were washed, and 100 μL of HRP-conjugated anti-human antibody was added to each well, followed by another half an hour incubation at 37°C. After five rounds of washing, 100 μL of chromogen-substrate was added to each well and incubated for 15 minutes. To halt the reaction, 100 μL of stop solution was added, and the absorbance was measured at 450 nm and 630 nm using a Hiperion Microplate Reader (MP4) instrument. After performing calculations using the original data and adjusting for dilution, the results were unveiled, shown as RU/mL antibody concentration.

### SARS-CoV-2 Anti spike IgG

To measure the amount of anti-spike IgG in serum samples, we utilized SARS-CoV-2 Spike (trimer) pre-coated plates (Thermo Fisher scientific, Lot: 359554-008) following the manufacturer’s instructions. Initially, the plates were washed twice, and then 100 μL of pre-diluted serum samples, controls, and standard solutions were added to the respective wells. The plates were incubated at 37°C for 30 minutes to allow reactions to happen. Subsequently, after another round of washing, 100 μL of HRP conjugate solution was added to each well. The plates were covered and were left at 37°C for another 30 minutes. Followed by three rounds of wash, each well received 100 μL of substrate solution. After 15 minutes of incubation at the room temperature, the process was stopped by addition of 100 μL of stop solution. The absorbance was measured at 450 nm using Hiperion Microplate Reader (MP4) immediately after adding the stop solution. The concentration of samples (U/mL) was calculated by multiplying the obtained value(s) for the sample(s) by the appropriate correction factor to account for sample dilution.

### Neutralization Assays

#### Pseudovirus neutralization assay (pVNT)

To generate Spike pseudo-typed lentivirus, plex307-egfp, pCMV3-spike (Wuhan-D614G), and pspax2 were co-transfected using Lipofectamine 3000 from Thermo-Scientific kit. The Spike pseudo-typed lentivirus carried the EGFP protein as a reporter gene. The evaluation of pVNT was executed in accordance with the procedures previously delineated ^33^. In brief, serum samples were diluted in a 2-fold serial manner, starting from 1/20, and dispensed into a 96-well cell culture plate. Simultaneously 50μL Spike pseudo-typed lentivirus was added to each well, and then the plate was incubated one hour at 37°C. Afterwards, the wells were supplemented with 14×10^3^ HEK-293T-hACE2 cells. After an incubation period of 48 to 60 hours, we determined the number of EGFP-positive cells in each well using fluorescent microscopy. Finally, The IC_50_ value was determined using the percentage of GFP-positive cells plotted against the logarithm of the dilution factor.

### Conventional virus neutralization test (cVNT)

In this study, the cVNT analysis was carried out using serum samples gathered both prior to vaccination and 14 days post-vaccination (study day 15). To conduct the assay, 100 μL of SARS-CoV-2 at a concentration of 10^2^ median cell culture infectious dose 50% per mL (CCID_50_/mL) was blended with 100 μL of heat-inactivated serum, which had been previously diluted in a two-fold serial manner. After a 60-minute incubation at 37°C, the mixture of virus and serum was introduced into a 96-well plate containing Vero cells, which had been seeded with a density of 1.5 × 10^4^ cells per well. This procedure was replicated three times. Subsequent to an hour of incubation, the supernatant was withdrawn, and the cells were washed twice using Dulbecco’s Modified Eagle Medium (DMEM). Following that, the cells were kept at 37°C in a 5% CO_2_ environment, maintained in DMEM supplemented with 10% heat-inactivated Fetal Bovine Serum (FBS) for a span of 72 hours. To determine IC_50_ titer, microscopic observation was employed. The most diluted solution that effectively prevented the occurrence of 50% cytopathic effect (CPE) was determined and reported as the IC_50_ titer.

### Intracellular cytokine staining

Intracellular cytokine staining followed by flow cytometry analysis was used to identify Spike-specific T cell responses. In short, previously cryopreserved peripheral blood mononuclear cells (PBMCs) were thawed, washed, and allowed to rest in RPMI 1640 (Biowest™) containing 10% heat inactivated FBS (Gibco™) supplemented with 2 μg/ml DNase I (Kiagene Fanavar), for 6-8 hours prior to stimulation. Afterwards, PBMCs were plated at approximately 2 × 10^6^ cell per well in a 96-well plate and restimulated with SARS-CoV-2 peptide pool (JPT Peptide Technologies) at concentration of 2 μg/mL of each peptide. PMA/Ionomycin (BD Pharmingen™) was utilized as a positive control and dimethyl sulfoxide (DMSO) (Sigma–Aldrich™) served as negative control agent. Brefeldin-A (Golgi-stop, BioLegend) was added to each well after two hours of incubation. After a total of 16 hours of stimulation at 37°C, the cells were collected and washed with phosphate-buffered saline (PBS) and then stained with Zombie Violet dye (BioLegend) at a dilution of 1/500. This staining step was performed at room temperature for a duration of 30 minutes. Then the stained cells were washed with FACS buffer (PBS supplemented with 2% FBS and 0.05% NaN3). Afterwards, the anti-CD3 antibody conjugated with Alexa Fluor^®^ 700 (BioLegend) was applied for surface staining. This was followed by an incubation of 20 minutes on ice. In order to stain intracellularly, samples were then fixed and permeabilized using a Cytofast fix/perm buffer set (BioLegend) as per the manufacturer’s instructions. Subsequently, a panel of antibodies, all sourced from BioLegend, including APC/Fire™ 810-CD4, PE/Fire™ 700-CD8, APC-IL-4, and Brilliant Violet 510™-IFN-γ, were applied and incubated for 20 minutes at room temperature. After a round of washing with permeabilization and staining buffer, the cells were subjected to analysis using a flow cytometer (BD FACS Lyrics) and the data were analyzed using FlowJo V10 software (BD Biosciences). Cytokine positive cells were identified by gating on lymphocytes, singlets, viability dye-, CD3^+^ followed by CD4^+^ or CD8^+^, displayed in supplementary figure 1. Unstimulated samples from the same individuals were used for determining the positive gates.

### Statistical analysis

In order to make the graphical representation and statistical analysis of the results, the software GraphPad V8 Prism was used. As a means to compare two groups of values, a t-test was performed, and a one-way ANOVA followed by Dunn’s test was used to compare results of multiple groups. A p-value of less than 0.05 was considered statistically significant (*P<0.05, **P<0.01, ***P<0.001, ****P<0.0001).

## ACKNOWLEDGEMENTS

We acknowledge the Iran National Innovation Fund (INIF) for financial support for this study, and Ministry of Health and Medical Education, Iran Food and Drug Administration (IFDA), Iran National Committee for Ethics in Biomedical Research, and Vice-presidency for Science and Technology (VPST), for all their support for this clinical trial. The authors would like to thank Professor Hossein Ghanaati, president of the Tehran University of Medical Sciences, for his valuable support, and the dedicated nurses of the Mahdi Clinic of the Imam Khomeini Hospital Complex (Tehran, Iran), where this study was conducted and all clinical trial coordinators. We sincerely thank all the volunteers who generously participated in this study. We would also like to thank the Noor Pathobiology and Genetic Laboratory and the Amirabad Laboratory for masked laboratory assessment of clinical samples of this study. We appreciate efforts of the members of the Data and Safety Monitoring Board (DSMB) for their careful evaluation of the clinical trial outcomes. We are also very grateful for the collaboration with AryoGen Pharmed, which contributed to the GMP-compliant manufacturing of COReNAPCIN^®^, and Pharmed Pajoohan Viera, as the contract research organization (CRO), for their participation in the design and conduct of this clinical trial. We also acknowledge all staff members of ReNAP Therapeutics who participated in logistics, procurement, production, quality assurance and quality control of COReNAPCIN^®^ and Placebo vials for this study.

## AUTHOR CONTRIBUTIONS

The authors M.S., I.A.D., H.H., M.A. S.M.H., V.K. and S.D. designed the clinical trial study. M.S, I.A.D., N.K. and M.A. managed and conducted and A.N. coordinated the clinical trial. M.A. S.P. and R.A under the supervision S.M.H. participated in masked immunological assessment of clinical samples. M.P., M.S. & M.M.S. participated in GMP-compliant manufacturing and quality assessment of COReNAPCIN^®^ and Placebo vials. A.N., V.K., M.A., S.P. M.S. and S.D wrote the manuscript. A.N., V.K., M.A, M.A., S.P. and S.D. made the figures and tables. S.D., S.M.H., and H.H. edited the manuscript.

## DATA AVAILABILITY

The data that support the findings of this study are available upon request from corresponding author Vahid Khoddami by email.

