## Supplementary File for "Safety and immunogenicity of COReNAPCIN^®^, a SARS-CoV-2 mRNA vaccine; a randomized, double-blind, placebo-controlled phase 1 clinical trial"

*[1]Research center for antibiotic stewardship and antimicrobial resistance, Imam Khomeini hospital complex, Tehran University of Medical Sciences, Tehran, Iran, [2]Department of Infectious Diseases, Loghman Hakim Hospital, Shahid Beheshti University of Medical Sciences, Tehran, Iran, [3]Infectious Diseases and Tropical Medicine Research Center, Shahid Beheshti University of Medical Sciences, Tehran, Iran, [4]ReNAP Therapeutics, Tehran, Iran, [5]Department of Internal Medicine, School of Medicine, Tehran University of Medical Sciences, Tehran, Iran, [6]Department of Medical Affairs, Pharmed Pajoohan Viera, Tehran, Iran, [7]Clinical Trial Center, Tehran University of Medical Sciences, Tehran, Iran, [8]Department of Immunology, School of Medicine, Shahid Beheshti University of Medical Sciences, Tehran, Iran, [9]Tehran University of Medical Sciences, Tehran, Iran.*

### Supplementary Datasets

### Supplementary Datasets

**Table 1. Summary of solicited Treatment Adverse Events According to Dose Groups.**

| Adverse Event | Group | Severity | | |
|---|---|---|---|---|
| | | Mild Count (N) | Moderate Count (N) | Severe Count (N) |
| Injection Site Pain | 50 µg | 4 | 7 | 1 |
| | 25 µg | 7 | 3 | 2 |
| | Placebo | 2 | 0 | 0 |
| Injection Site Erythema | 50 µg | 1 | 0 | 0 |
| | 25 µg | 4 | 0 | 0 |
| | Placebo | 0 | 0 | 0 |
| Injection site Edema/Induration | 50 µg | 1 | 0 | 0 |
| | 25 µg | 4 | 1 | 0 |
| | Placebo | 1 | 0 | 0 |
| Headache | 50 µg | 2 | 3 | 0 |
| | 25 µg | 1 | 1 | 2 |
| | Placebo | 1 | 1 | 0 |
| Fatigue | 50 µg | 3 | 5 | 1 |
| | 25 µg | 3 | 2 | 0 |
| | Placebo | 2 | 0 | 0 |
| Myalgia | 50 µg | 5 | 5 | 1 |
| | 25 µg | 2 | 0 | 1 |
| | Placebo | 0 | 1 | 0 |
| Arthralgia | 50 µg | 2 | 1 | 0 |
| | 25 µg | 0 | 0 | 0 |
| | Placebo | 1 | 1 | 0 |
| Fever | 50 µg | 6 | 0 | 0 |
| | 25 µg | 2 | 0 | 0 |
| | Placebo | 0 | 0 | 1 |
| Chills | 50 µg | 3 | 2 | 1 |
| | 25 µg | 1 | 0 | 0 |
| | Placebo | 0 | 1 | 0 |
| Nausea/vomiting | 50 µg | 0 | 0 | 0 |
| | 25 µg | 0 | 0 | 0 |
| | Placebo | 0 | 0 | 0 |
| Diarrhea | 50 µg | 1 | 0 | 0 |
| | 25 µg | 2 | 0 | 0 |
| | Placebo | 0 | 0 | 0 |
| Axillary Lymphadenopathy | 50 µg | 0 | 1 | 0 |
| | 25 µg | 2 | 0 | 0 |
| | Placebo | 0 | 0 | 0 |

**Table 2. Summary of Unsolicited Treatment Adverse Events According to Dose Groups.**

| Adverse Event | Group | Severity | | |
|---|---|---|---|---|
| | | Mild Count (N) | Moderate Count (N) | Severe Count (N) |
| Abdominal cramps | 50 µg | 1 | 1 | 0 |
| | 25 µg | 2 | 0 | 0 |
| | Placebo | 0 | 0 | 0 |
| Allergic skin reaction | 50 µg | 0 | 0 | 0 |
| | 25 µg | 0 | 0 | 0 |
| | Placebo | 1 | 0 | 0 |
| Back pain | 50 µg | 0 | 0 | 0 |
| | 25 µg | 0 | 0 | 0 |
| | Placebo | 0 | 0 | 1 |
| Blood pressure increased | 50 µg | 0 | 1 | 0 |
| | 25 µg | 0 | 0 | 0 |
| | Placebo | 0 | 0 | 0 |
| Canker sore lip | 50 µg | 0 | 1 | 0 |
| | 25 µg | 0 | 0 | 0 |
| | Placebo | 0 | 0 | 0 |
| Chest heaviness | 50 µg | 0 | 1 | 0 |
| | 25 µg | 0 | 0 | 0 |
| | Placebo | 0 | 0 | 0 |
| Chest pain | 50 µg | 0 | 0 | 0 |
| | 25 µg | 0 | 0 | 0 |
| | Placebo | 0 | 0 | 1 |
| Common cold | 50 µg | 0 | 2 | 0 |
| | 25 µg | 1 | 3 | 1 |
| | Placebo | 0 | 1 | 0 |
| Corona virus infection | 50 µg | 0 | 0 | 0 |
| | 25 µg | 0 | 0 | 0 |
| | Placebo | 0 | 1 | 0 |
| Dental and periodontal infections and inflammations | 50 µg | 0 | 0 | 0 |
| | 25 µg | 0 | 0 | 0 |
| | Placebo | 0 | 1 | 0 |
| Diarrhea | 50 µg | 0 | 0 | 1 |
| | 25 µg | 1 | 1 | 0 |
| | Placebo | 0 | 0 | 0 |
| Dizziness | 50 µg | 0 | 2 | 0 |
| | 25 µg | 0 | 1 | 0 |
| | Placebo | 0 | 0 | 0 |
| Dry cough | 50 µg | 0 | 0 | 0 |
| | 25 µg | 0 | 1 | 0 |
| | Placebo | 0 | 0 | 0 |
| Eczema exacerbated | 50 µg | 0 | 0 | 0 |
| | 25 µg | 1 | 0 | 0 |
| | Placebo | 0 | 0 | 0 |

| | | | | |
|---|---|---|---|---|
| Eye Pain | **50 µg** | **0** | **0** | **0** |
| | **25 µg** | **1** | **0** | **1** |
| | **Placebo** | **0** | **0** | **0** |
| Eyes tearing | **50 µg** | **0** | **1** | **0** |
| | **25 µg** | **0** | **0** | **0** |
| | **Placebo** | **0** | **0** | **0** |
| Fatigue | **50 µg** | **1** | **0** | **0** |
| | **25 µg** | **0** | **0** | **0** |
| | **Placebo** | **0** | **0** | **0** |
| Feeling hot | **50 µg** | **0** | **0** | **0** |
| | **25 µg** | **2** | **0** | **0** |
| | **Placebo** | **0** | **0** | **0** |
| Fever | **50 µg** | **1** | **0** | **0** |
| | **25 µg** | **0** | **0** | **0** |
| | **Placebo** | **0** | **0** | **0** |
| Hand pain | **50 µg** | **0** | **1** | **0** |
| | **25 µg** | **0** | **0** | **0** |
| | **Placebo** | **0** | **0** | **0** |
| Headache | **50 µg** | **0** | **5** | **0** |
| | **25 µg** | **2** | **1** | **0** |
| | **Placebo** | **1** | **0** | **0** |
| Heart rate increased | **50 µg** | **0** | **1** | **0** |
| | **25 µg** | **0** | **0** | **0** |
| | **Placebo** | **0** | **0** | **0** |
| Heartburn | **50 µg** | **0** | **0** | **0** |
| | **25 µg** | **1** | **0** | **0** |
| | **Placebo** | **0** | **0** | **0** |
| Hiccups | **50 µg** | **0** | **1** | **0** |
| | **25 µg** | **0** | **0** | **0** |
| | **Placebo** | **0** | **0** | **0** |
| Hoarseness of voice | **50 µg** | **0** | **0** | **0** |
| | **25 µg** | **0** | **1** | **0** |
| | **Placebo** | **0** | **0** | **0** |
| Injection site bruising | **50 µg** | **0** | **0** | **0** |
| | **25 µg** | **0** | **0** | **0** |
| | **Placebo** | **1** | **0** | **0** |
| Injection site tingling | **50 µg** | **0** | **0** | **0** |
| | **25 µg** | **1** | **0** | **0** |
| | **Placebo** | **0** | **0** | **0** |
| Injection site warmth | **50 µg** | **0** | **0** | **0** |
| | **25 µg** | **1** | **0** | **0** |
| | **Placebo** | **0** | **0** | **0** |
| Itchy throat | **50 µg** | **0** | **1** | **0** |
| | **25 µg** | **0** | **0** | **0** |
| | **Placebo** | **0** | **0** | **0** |
| Myalgia | **50 µg** | **0** | **0** | **0** |
| | **25 µg** | **0** | **0** | **0** |

| | | | | |
|---|---|---|---|---|
| | **Placebo** | **1** | **0** | **0** |
| Neck pain | **50 µg** | **0** | **0** | **0** |
| | **25 µg** | **0** | **0** | **0** |
| | **Placebo** | **1** | **0** | **0** |
| Paresthesia of fingers | **50 µg** | **1** | **0** | **0** |
| | **25 µg** | **0** | **0** | **0** |
| | **Placebo** | **0** | **0** | **0** |
| Restless legs | **50 µg** | **0** | **0** | **0** |
| | **25 µg** | **1** | **0** | **0** |
| | **Placebo** | **0** | **0** | **0** |
| Runny nose | **50 µg** | **1** | **0** | **0** |
| | **25 µg** | **0** | **0** | **0** |
| | **Placebo** | **0** | **0** | **0** |
| Seasonal allergic rhinitis | **50 µg** | **1** | **0** | **0** |
| | **25 µg** | **0** | **0** | **0** |
| | **Placebo** | **0** | **0** | **0** |
| Skin injury | **50 µg** | **0** | **1** | **0** |
| | **25 µg** | **0** | **0** | **0** |
| | **Placebo** | **0** | **0** | **0** |
| Stomachache | **50 µg** | **1** | **0** | **0** |
| | **25 µg** | **0** | **0** | **0** |
| | **Placebo** | **0** | **0** | **0** |
| Temporal headache | **50 µg** | **0** | **0** | **0** |
| | **25 µg** | **0** | **0** | **0** |
| | **Placebo** | **1** | **0** | **0** |
| Tingling feet/hands | **50 µg** | **0** | **0** | **0** |
| | **25 µg** | **2** | **0** | **0** |
| | **Placebo** | **0** | **0** | **0** |
| Tiredness | **50 µg** | **0** | **0** | **0** |
| | **25 µg** | **0** | **1** | **0** |
| | **Placebo** | **0** | **0** | **0** |
| Weakness | **50 µg** | **0** | **0** | **0** |
| | **25 µg** | **1** | **0** | **0** |
| | **Placebo** | **0** | **0** | **0** |

**Table 3. Summary of Anti-spike concentration in 25 μg, 50 μg and Placebo groups.**

| Group \ Day | Day 1 | Day 8 | Day 15 | Day 29 |
|---|---|---|---|---|
| | **Anti-spike IgG sero-conversion Geomean** (Lower-Upper 95% CI of geo. Mean) | | | |
| 25 μg | **13477** (6898-26332) | **33640** (15306-73932) | **164322** (110773-243756) | **104123** (74980-144593) |
| 50 μg | **10488** (5164-21301) | **82192** (43600-154941) | **227954** (137345-378341) | **159605** (97496-261281) |
| placebo | **10333** (3326-32104) | **8960** (4013-20006) | **12459** (3196-48563) | **10860** (4635-25447) |

**Table 4. Summary of Anti-RBD concentration in 25 μg, 50 μg and Placebo groups.**

| Group \ Day | Day 1 | Day 8 | Day 15 | Day 29 |
|---|---|---|---|---|
| | **Anti-RBD IgG sero-conversion Geomean** (Lower-Upper 95% CI of geo. Mean) | | | |
| 25 μg | **52.90** (30.24- 92.54) | **127.3** (67.56-239.8) | **359.0** (206.0-625.6) | **314.0** (197.2-500.1) |
| 50 μg | **54.72** (27.52-108.8) | **328.4** (199.5-540.6) | **443.4** (282.5-695.9) | **393.1** (241.7-639.6) |
| placebo | **39.02** (12.99-117.3) | **37.72** (13.07-108.9) | **44.95** (8.119- 248.9) | **51.80** (18.20-147.4) |

**Supplementary Figure 1. Gating strategy used for identification of SARS-CoV-2–reactive T cells.**

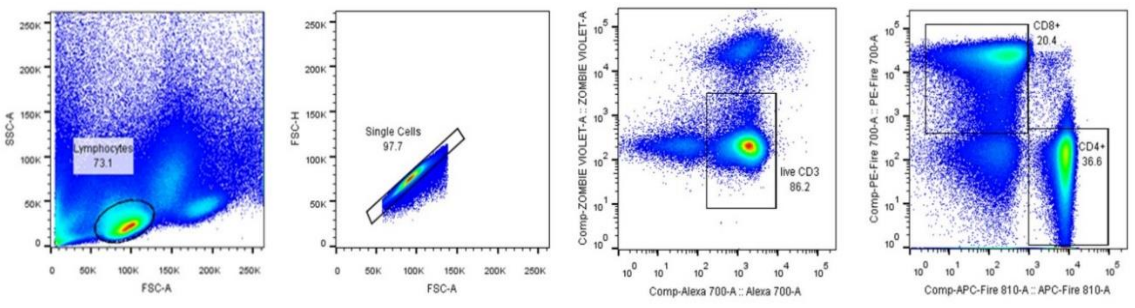
